# Evaluating the accuracy of self-collected swabs for the diagnosis of monkeypox

**DOI:** 10.1101/2022.09.19.22280087

**Authors:** Maria Ubals, Eloy José Tarín-Vicente, Xènia Oller, Adrià Mendoza, Andrea Alemany, Águeda Hernández-Rodríguez, Cristina Casañ, Ángel Rivero, Pep Coll, José Miguel Cabrera, Martí Vall, Manuel Agud-Dios, Elena Gil-Cruz, Alexia Paris de Leon, Aída Ramírez, María Dolores Folgueira, María Angeles Melendez, Vira Buhiichyk, Cristina Galván-Casas, Roger Paredes, Nuria Prat, Maria-Rosa Sala Farre, Josep Maria Bonet-Simó, Pablo L Ortiz-Romero, Bonaventura Clotet, Pere-Joan Cardona, Ignacio Blanco, Michael Marks, Clara Suñer, Oriol Mitjà, The Movie Group

## Abstract

We evaluated the accuracy of patient-collected skin lesions, pharyngeal, and rectal swabs amongst 50 individuals enrolled in a study of monkeypox viral dynamics. We found that the performance of self-collected samples was similar to that of physician-collected samples, suggesting that self-sampling is a reliable strategy for diagnosing monkeypox.

## Background

In early 2022, an outbreak of monkeypox was reported in Europe, and cases have subsequently been reported worldwide [1]. Unlike previous monkeypox outbreaks, individuals infected during 2022 have been predominantly men who have sex with men (MSM) without any history of travel to an endemic country. Based on clinical and risk factor data, transmission during sex appears to be the primary driver of the current epidemic, and patients typically present with a high frequency of skin lesions in the genital, peri-anal, and perioral region and proctitis, tonsilitis, and penile oedema as the most common complications [2–4].

Diagnosis of monkeypox relies on detecting viral DNA by PCR testing [5] in several body specimens. In the current outbreak, the highest yield has been reported in samples collected from skin lesions, although the virus is also frequently detectable in throat and rectal swabs, as well as in blood [3,6]. Detection in urine and semen have been reported in some studies but these samples are not routinely used for diagnosis [7].

Self-sampling is a strategy where the patient ―and not a healthcare provider― collects the clinical samples required for diagnosis. This strategy has been well established for the diagnosis of many sexually transmitted infections, with similar performance to samples collected by physicians [8,9]. More recently, self-sampling has been shown to be a reliable strategy for detecting SARS-CoV-2 infection [10]. To test the performance of this strategy in the monkeypox setting, we nested an evaluation of self-sampling within a more extensive study on the viral dynamics of monkeypox.

## Methods

### PARTICIPANTS

We conducted a prospective diagnostic accuracy evaluation in individuals with suspected monkeypox in three centres in Spain. All patients presenting to participating centres with lesions suggestive of monkeypox and compatible symptoms starting within the 10 days preceding screening underwent a standardised clinical assessment by a dermatologist or a specialist in sexually transmitted infections and were invited to participate. Patients who required hospital admission were not included in the study.

### PROCEDURES

At the baseline visit, a physician collected clinical samples, including lesion, pharyngeal, and rectal swabs as appropriate (study day 0). Participants were provided with home testing kit materials, which included an instruction sheet and devices for self-collection (i.e., Dacron-tipped swabs, pre-labelled swab containers, and a mailing envelope); they were trained for self-collection of samples and asked to self-collect swabs from the same skin lesions, the oropharynx, and the rectum the following day (study day 1). Participants were instructed to keep samples at 4 ºC after collection and contact the parcel courier service, which transferred the samples to the microbiology laboratory of the University Hospital Germans Trias i Pujol (Badalona, Spain). Swabs were analysed using a quantitative polymerase chain reaction (qPCR), as described elsewhere. Briefly, monkeypox virus DNA was detected by LightMix Modular Orthopox Virus assay (TIB MolBiol, Berlin, Germany) on LightCycler 480 Real-Time PCR equipment (Roche Applied Science, Mannheim, Germany) by amplifying a 113-base-pair-long fragment gene specific to orthopoxviruses [5]. Patients with a positive result in any of the samples collected by a physician on day 0 were classified as having monkeypox virus and, therefore, included in the study analyses.

### STATISTICAL ANALYSIS

Continuous and categorical variables are presented as the median and interquartile range [IQR], defined by 25th and 75th percentiles) and number (%), respectively. We considered that discrepant results might occur in either direction (i.e., a physician-collected swab might be negative and a self-collected swab positive or vice versa) because of sampling error. Therefore, we calculated the overall agreement between self-collected swabs on study day 1 (index test) and physician-collected swabs on study day 0 (reference test) using the kappa statistic. In a secondary analysis, we compared the viral load, as measured by the cycle threshold (CT) value, between physician- and self-collected samples using a paired t-test. All analyses were conducted in R version 4.2.1. The cut-off CT for real-time amplification assays was 40, above which the sample was considered negative.

### ETHICAL APPROVAL

The study was approved by the Ethics Committee of the Hospital Germans Trias i Pujol. Written informed consent was obtained from all participants.

## Results

We enrolled 50 patients with suspected monkeypox. All the patients were male, and the median age was 33.5 years (IQR 28-45.5 years). All patients had PCR-confirmed monkeypox in at least one of the diagnostic specimens collected. The median time from symptoms onset was 5 days (IQR 4-6 days). At baseline, 49 individuals had a skin lesion swab collected (all of which were positive), 38 had a pharyngeal swab collected (26 [68%] were positive), and 11 had a rectal swab collected (9 [82%] were positive).

Paired samples were available in 49, 38, and 10 individuals for skin lesions, throat, and rectal specimens, respectively (Figure 1). The highest overall agreement was observed in lesional skin swabs (98% agreement), where only one individual tested negative in the physician-collected swab and positive in the self-collected. For throat and rectal specimens, the overall agreement was 79% (kappa 0.49) and 90% (kappa 0.6), respectively.

**Figure 1:**
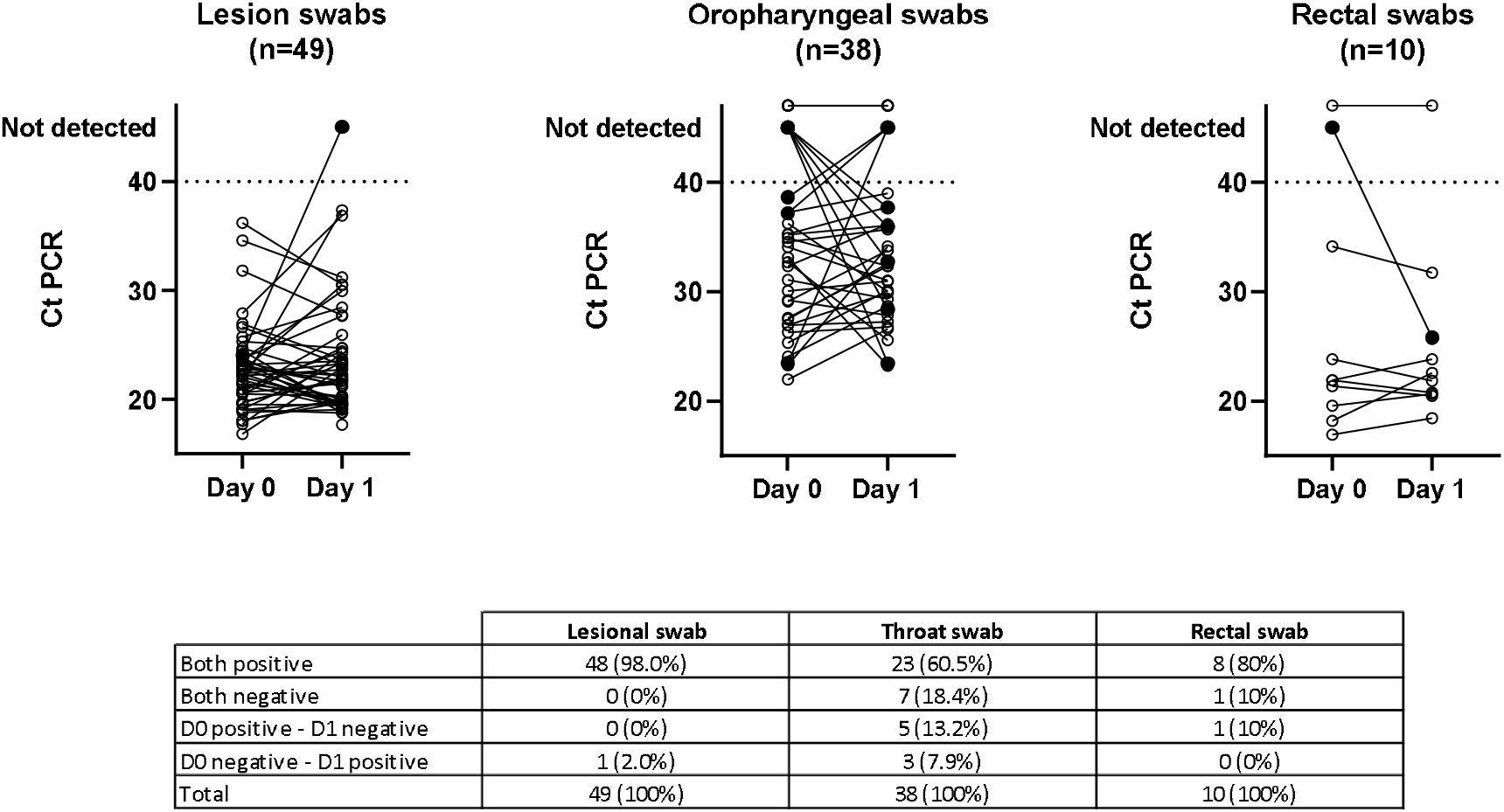
Concordance and discordance between physician and self-collected swabs. Cycle threshold (CT) values of swabs collected by the physician on day 0 compared to self-collection by the patient on Day 1. Concordant results are showed as clear circles. Discordant results are coloured black. The Ct value of one skin lesion swab positive on day 0 was not available and this sample and its corresponding day 1 sample were therefore not included in the figure.

We found no significant differences in CT values between physician- and self-collected skin lesion and throat specimens. The mean CT values of physician- and self-collected lesion swabs were 22.5 and 23.2, respectively (absolute difference 0.7; 95% CI −2.5 to 1.1, *p* = 0.44). The mean CT values for physician- and self-collected oropharyngeal swabs were 30.6 and 30.7, respectively (absolute difference 0.1; 95% CI −2.6 to −2.3, *p* = 0.92). Conversely, for rectal swabs, self-collected samples had higher CT values than physician-collected samples (absolute difference 5.5; 95% CI −10 to −1.0, *p* <0.02).

## Discussion

This is the first study to demonstrate the feasibility and accuracy of self-collected samples for the diagnosis of monkeypox. Overall, self-collected swabs had high accuracy and similar viral loads to physician-collected swabs. Self-sampling for the diagnosis of sexually transmitted infections utilising self-taken pharyngeal, genital, and rectal swabs is a well-established strategy for diagnosing chlamydia and gonorrhoea based on nucleic acid amplification testing [8,9]. Our data extend these findings to confirm the applicability of this strategy in monkeypox. Importantly, we show patient-collected samples from skin lesions to have similarly high-performance characteristics. Unlike the other types of samples, patient-collected skin swabs are not routinely used to diagnose common blistering skin diseases such as herpes or varicella, but our data demonstrate the applicability of this approach.

The agreement between physician- and self-taken oropharyngeal swabs was lower than for other samples. Oropharyngeal swabs are likely more prone to variation in the quality of sample collection compared to easily visualized skin lesions or rectal swabs. We believe that differences in sampling most likely explain these discordant results; however, fluctuations in viral load within the pharynx are also possible.

Our study has some limitations. Firstly, we enrolled a limited number of participants. Ideally, a larger sample size would allow greater precision in our estimates of accuracy. However, our findings are consistent with a considerable burden of literature on self-sampling; therefore, it seems unlikely that a larger sample size would fundamentally alter our findings. Secondly, the type of samples taken on day 0 was at physician’s discretion before patient enrolment. Consequently, some patients without proctitis lacked a physician-collected sample, resulting in fewer paired rectal samples than in other locations. Nevertheless, our data are consistent with existing literature indicating a similar performance of self-collected and physician-collected rectal samples. Finally, samples for the reference and index tests were taken 1 day apart. Although test performance studies are typically cross-sectional, we considered that samples taken at home without professional support or supervision would provide a more accurate view of the diagnostic performance based on self-sampling. Samples testing positive at baseline and negative on day 1 could reflect a true negative due to clearance of the virus rather than inadequate sampling. However, we equally noted that some samples, in particular oropharyngeal swabs collected on day 1, tested positive despite a negative result on day 0. Considering that the viral loads were very similar at both time points, it seems most likely that these changes represent variation in sampling rather than real changes in viral load.

Our data confirm that a variety of self-taken samples can be used to reliably diagnose monkeypox in individuals with suspected signs of monkeypox infection. The self-sampling strategy offers a number of potential advantages for patients and disease control and facilitates the integration of monkeypox into routine testing for other sexually transmitted infections in high-risk populations. Further work to optimize sample collection, including consideration of other types of samples, such as saliva, could be considered to further enhance the ease of testing.

## Data Availability

All data produced in the present study are available upon reasonable request to the authors

## Acknowledgements

We thank Gerard Carot-Sans for editing the final draft and Roser Escrig for medical writing assistance with the study documentation. We also thank Laia Bertran, Sergi Gavilan, and Miquel Angel Rodríguez for the operational and financial management of the project. OM, CS, MU and AA are supported by the European Research Council grant agreement number 850450 (European Union’s Horizon 2020 Research and Innovation Program, ERC-2019-STG funding scheme). This work was funded by the JoEmCorono crowdfunding campaign.

